# SPATIAL ANALYSIS OF EXCLUSIVE BREASTFEEDING COVERAGE AND SOCIO-HEALTH DETERMINANTS IN MUARAENIM REGENCY, INDONESIA

**DOI:** 10.64898/2026.01.12.26343922

**Authors:** Yona Wia Sartika Sari, Prihatini Dini Novitasari, Alvera Noviyani, Ayu Prameswari

## Abstract

**Background:** Exclusive breastfeeding (EBF) for the first six months is a cornerstone of child survival, yet its coverage remains uneven across regions in Indonesia.

**Methods:** This study examined the spatial distribution of EBF coverage alongside maternal and service-related characteristics in Muara Enim Regency using a quantitative descriptive spatial approach. Secondary data from 1,267 mothers with children under five years of age in the Gelumbang, Lembak, and Kelekar sub-districts were analyzed using QGIS. The variables included EBF coverage, early initiation of breastfeeding (EIBF) coverage, maternal employment status, maternal education level, and place of delivery. Spatial visualization was conducted through pie diagram mapping and overlay techniques at the village and sub-district levels.

**Results:** The findings revealed clear geographic disparities in EBF coverage. Spatial overlays showed that differences in EIBF practices, maternal education, employment status, and place of delivery were not consistently aligned with areas of higher EBF coverage.

**Conclusion:** These findings underscore the relevance of incorporating spatial perspectives into public health planning to address disparities in breastfeeding practices.

## INTRODUCTION

Exclusive breastfeeding (EBF) is the feeding of only infants after the baby is born until 6 months of age, and there are no liquids or other foods except for a few drops of syrup consisting of vitamins, mineral supplements, oral rehydration saline solution (ORS), or medications as medically indicated.^1^ EBF is recommended by the World Health Organization (WHO) for infants during the first six months of life due to its complete nutritional content and its role in supporting the child’s immune system.^2^ The Indonesian government has targeted an increase in exclusive breastfeeding coverage nationally, but the target still shows inequality between regions. Data from the Ministry of Health of the Republic of Indonesia indicate that the percentages of infants who received exclusive breastfeeding in the 2020-2023 period were 66%, 56.9%, 61.5%, and 63.9%, respectively. This achievement has exceeded the national program target for 2023 of 50%.^3–5^ Meanwhile, EBF coverage in South Sumatra is 69.22% in 2024, with a strategic plan target of 60%. This coverage increased by 4.72% compared with 2023, which was 64.5%.^6^

Muara Enim Regency, like many other regions in South Sumatra, exhibits diverse geographic and demographic characteristics that may contribute to health disparities. In South Sumatra, EBF coverage was 66.3% in 2022 and 68.9% in 2023. This is still below the program’s goal of 70%. Meanwhile, EBF coverage in Muara Enim is 71.1% in 2022 and 71.4% in 2023.^7,8^

The success of an EBF program depends not only on the mother’s individual factors but also on the social context and the accessibility of surrounding health services.^9^ Early Initiation of Breastfeeding (EIBF) is a highly beneficial factor for the sustainability of EBF and prolonged breastfeeding.^5^ Maternal education level is also an important determinant, with mothers with higher levels of education more likely to understand the benefits of EBF.^10^ In addition, the mother’s employment status is closely associated with exclusive breastfeeding practices.^11^ Other studies also report a significant association between work and EBF.^12^

The Government of Indonesia has made various efforts to increase the success of exclusive breastfeeding through health policies, regulations, and programs. One of the strategic steps is the implementation of Government Regulation Number 33 of 2012 on EBF, which states that exclusive breastfeeding is the provision of breast milk to infants from birth for six months, without adding to or replacing it with other foods or beverages (except drugs, vitamins, and minerals).^13^ Furthermore, Law No. 4 of 2024 on the Welfare of Mothers and Children in the First 1000 Days stipulates that working mothers are entitled to maternity leave of six months.^14^ This provision demonstrates the support mothers receive in providing EBF to their babies.

Several studies have shown that EBF plays a crucial role as a primary protective factor against stunting incidence in children.^15^ EBF has a positive impact on the baby’s health. Infants who were exclusively breastfed had a lower incidence of diseases such as diarrhea, otitis media, urinary tract infections, allergic diseases, pneumonia, and protein-energy malnutrition than infants who were not exclusively breastfed.^16^ The success of EBF is influenced by social and health determinants, including maternal education, employment, partner support, health worker support services, and maternal knowledge of EBF.^17,18^

However, most previous studies have analyzed these factors without considering spatial or geographical dimensions. In fact, the distribution of health problems is often not random but forms a specific pattern or cluster. The geospatial approach allows policymakers to see “where” problems occur and “why” they are concentrated in specific locations. By mapping the exclusive breastfeeding coverage and the determinants influencing it, we can identify vulnerable areas that require special attention.

## METHODS

### Study Design

This study employed a descriptive spatial design to map the coverage of exclusive breastfeeding (EBF) alongside selected maternal and service-related characteristics in Muara Enim Regency. The overlay technique was used to superimpose two or more variables on a single map according to village or sub-district areas, allowing identification of spatial patterns formed by combinations of variables.

### Study Area

The research was conducted in three sub-districts of Muara Enim Regency: Kelekar, Gelumbang, and Lembak. Muara Enim Regency is geographically located between 4° and 6° South Latitude and 104° and 106° East Longitude, featuring diverse topography with plateaus in the southwest and lowlands in the central and northern regions. Not all villages within the sub-districts were included due to limitations in available data.

### Study Population and Sample

The study population comprised mothers residing in the selected sub-districts. A total of 1,267 mothers were included in this study. Sampling was based on secondary data collected during the Field Learning Experience (FLE) by students of the Faculty of Public Health, Sriwijaya University, in 2024. Mothers with incomplete data were excluded from the analysis.

### Study Variables

The study variables included the coverage of exclusive breastfeeding (EBF), the coverage of early initiation of breastfeeding (EIBF), maternal employment status, maternal education level, and place of delivery. All variables were measured and presented as percentages at the village or sub-district level and used for spatial mapping and overlay analysis.

### Data Sources

All variables were obtained from secondary data collected during the Field Learning Experience (FLE) conducted in 2024. The secondary data were used to create spatial maps illustrating patterns of maternal and service-related characteristics in relation to EBF and EIBF coverage.

### Data Processing and Analysis

Data were processed and visualized using a spatial overlay technique, where two or more variables were superimposed on a single map according to village or sub-district boundaries. This method allows the identification of spatial relationships and patterns between breastfeeding coverage and maternal or service-related characteristics. Descriptive statistics, including percentages and frequencies, were used to summarize maternal characteristics and service-related variables.

## RESULTS

Figure 1 shows exclusive breastfeeding coverage in the Gelumbang, Lembak, and Kelekar sub-districts, classified as high (60%) and low (<60%). Based on figure 1, there are villages/sub-districts that have high exclusive breastfeeding coverage, namely Teluk Jaya at 66%, while 27 other villages/sub-districts have low exclusive breastfeeding coverage, namely Gumai (35%), Suka Jaya (40%), Bitis (24%), Suka Menang (20%), Talang Taling (7%), Gelumbang (43%), Midar (20%), Sigam (35%), Tambangan Kelekar (13%), Karang Endah (35%), Jambu (36%), Embacang Kelekar (3%), Pelempang (13%), Karang Endah Selatan (41%), Talang Nangka (5%), Alai (36%), Petanang (23%), Sungai Duren (43%), South Alai (11%), Tapus (48%), Lembak (19%), Tanjung Baru (13%), Kemang (27%), Lubuk Enau (51%), Menanti (35%), Menanti Selatan (5%), and New Sudan (32%).

**Figure 1.**
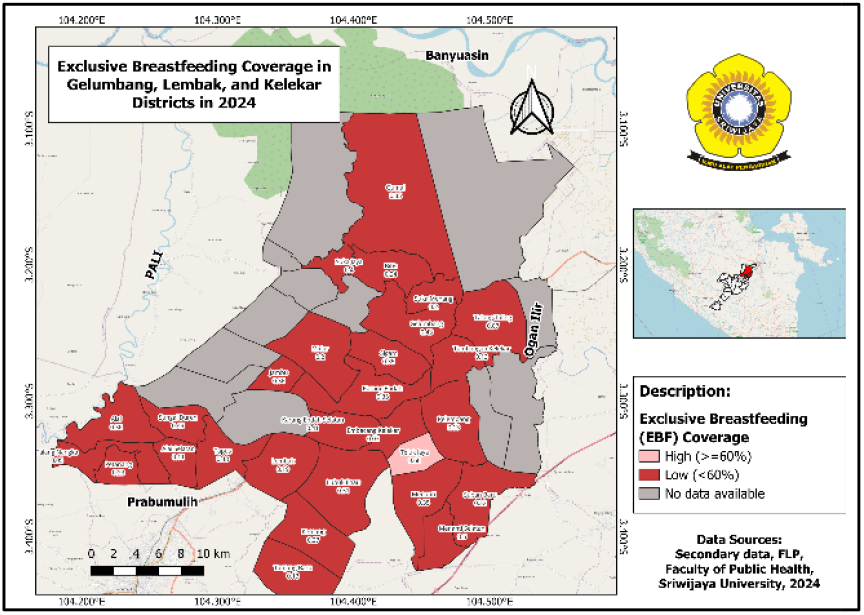
Map of Exclusive Breastfeeding Coverage by Village in Gelumbang, Lembak, and Kelekar Districts, 2024

Based on the mapping results, all villages/sub-districts in Gelumbang, Lembak, and Kelekar Districts showed high EIBF coverage, marked in yellow to indicate EIBF implementation (figure 2). However, EBF coverage shows that only the Teluk Jaya villages have high coverage ( ≥ 60%), whereas the other villages remain in the low category (<60%). This analysis shows that high EIBF coverage is not always accompanied by high EBF coverage; in this region, there is no direct relationship between EIBF implementation and EBF success.

**Figure 2.**
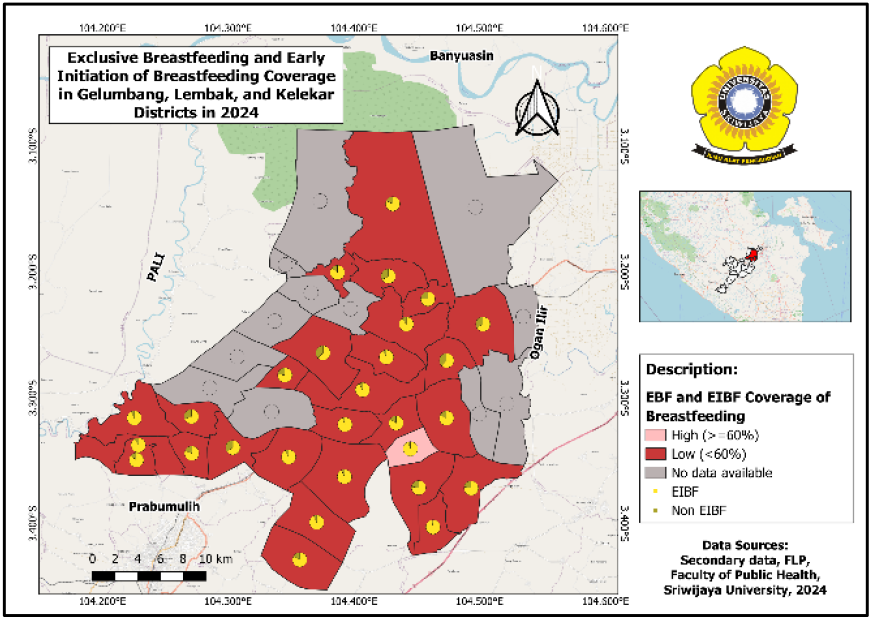
Map of Exclusive Breastfeeding and Early Initiation of Breastfeeding Coverage in Gelumbang, Lembak, and Kelekar Districts, 2024

Early initiation of breastfeeding within 1 hour of birth protects newborns from infection and reduces mortality. The risk of death from diarrhea and other infections may increase in babies who are partially breastfed or not breastfed at all.^19^

Based on the mapping results, all villages/sub-districts in the Gelumbang, Lembak, and Kelekar Districts are shown as light blue on the pie chart for each region (figure 3). Meanwhile, in terms of EBF coverage, the map shows that most areas are still in the low category (<60%), marked in red. This indicates that the high proportion of mothers who do not work is not fully accounted for by the increase in EBF coverage. This is in line with previous research in Indonesia, which states that many mothers only take care of household chores and do not exclusively breastfeed their babies.^20^

**Figure 3.**
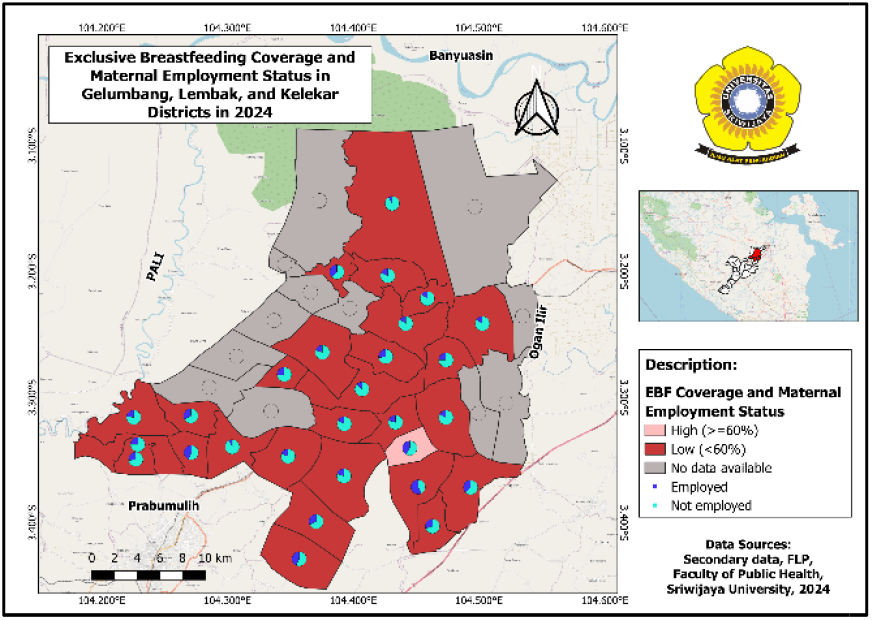
Map of Exclusive Breastfeeding Coverage and Maternal Employment Status in Gelumbang, Lembak, and Kelekar Districts, 2024

Based on the mapping results, all villages/sub-districts in Gelumbang, Lembak, and Kelekar sub-districts show that mothers who have higher education are as numerous as mothers with low education and are marked with light green and dark green colors on the pie diagram in each region (figure 4). Meanwhile, in terms of EBF coverage, the map shows that most areas are still in the low category (<60%), marked in red.

**Figure 4.**
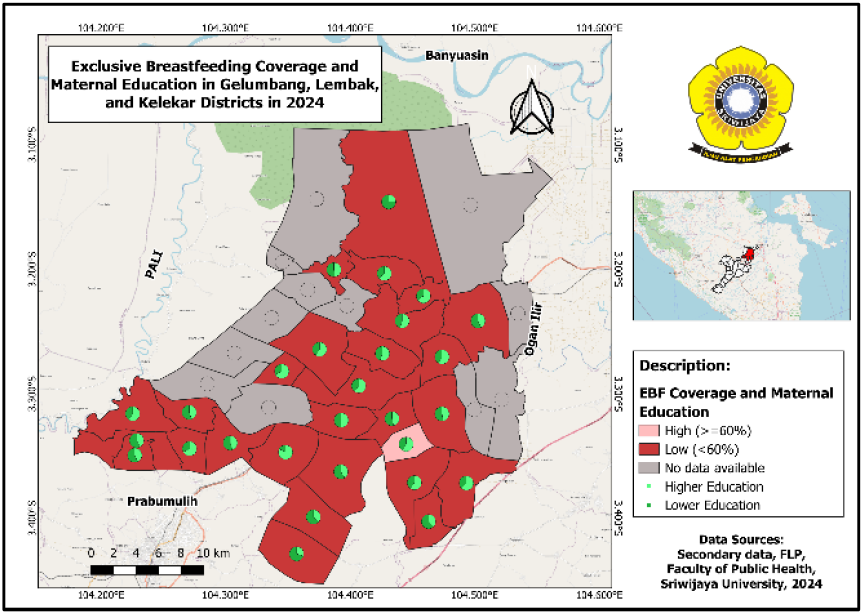
Map of Exclusive Breastfeeding Coverage and Maternal Education Level in Gelumbang, Lembak, and Kelekar Districts, 2024

Of the 14 villages with a proportion of highly educated mothers, only 1 has high exclusive breastfeeding coverage (60%), whereas the other 13 remain in the low category (<60%). This condition shows that the high level of maternal education is not fully proportional to the practice of exclusive breastfeeding in the region.

Based on the mapping results, all villages/sub-districts in the Gelumbang, Lembak, and Kelekar districts show that almost all mothers give birth in health service facilities, as indicated by the light purple in the pie diagrams for each region (figure 5). This illustrates that access to and utilization of maternal health services in the region are already relatively satisfactory.

**Figure 5.**
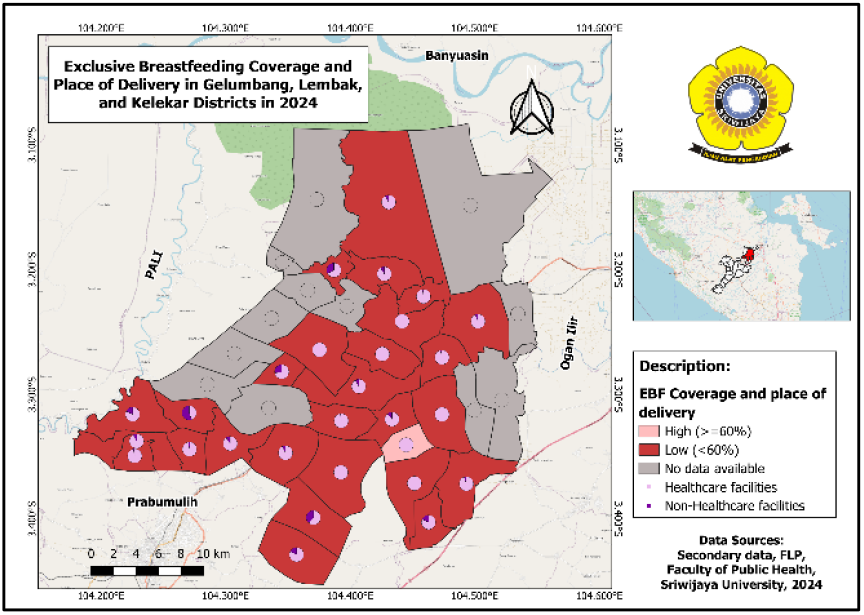
Map of Exclusive Breastfeeding Coverage and Place of Delivery in Gelumbang, Lembak, and Kelekar Districts, 2024

However, in terms of EBF coverage, the map shows that most of the region is still in the low category (<60%), which is marked in red. This condition shows that there is a gap between the behavior of using health services and the practice of EBF. Although mothers have given birth at health facilities, which should be the starting point for EBF promotion and education, breastfeeding practices are not optimally implemented.

## DISCUSSION

Based on the results of mapping exclusive breastfeeding coverage in villages/sub-districts in Gelumbang, Lembak, and Kelekar Districts, the proportion of mothers who give EBF to their babies is still low. EIBF factors, maternal employment status, maternal education, and place of delivery did not show an association in this study. There may be other factors influencing exclusive breastfeeding.

This study shows that areas with high EIBF coverage do not necessarily have high EBF coverage. In contrast to other studies conducted in Indonesia and Ethiopia, there is a significant association between a history of EIBF and exclusive breastfeeding practices.^21,22^ In contrast to studies in Spain, which reported no significant association between EIBF and EBF.^23^ The possibility of stopping breastfeeding before 6 months was associated with lactation counseling education by trained health workers, as well as family support, which made mothers more confident in EBF.^24^ Studies in Oman report that mothers who receive their husband’s support are twice as likely to breastfeed exclusively.^25,26^ Meanwhile, research in Indonesia shows that a husband’s positive attitude and involvement in helping his wife strengthen maternal motivation and help cope with various challenges during breastfeeding.^27^

The study shows that most mothers were not employed, while exclusive breastfeeding coverage remained low acros**s** the study areas. Research also conducted in Indonesia shows that employment status has a significant relationship with exclusive breastfeeding.^12^ Working mothers often have less time to breastfeed than mothers with more time to care for their babies at home. However, other studies have shown no association between maternal employment status and exclusive breastfeeding.^28^ It is possible that other obstacles prevent mothers from initiating exclusive breastfeeding, including local customs and cultural factors. Some cultures support exclusive breastfeeding, while others do not. Research in several rural areas of Indonesia shows that there is still a traditional practice of giving honey to newborns and giving solid food such as bananas or porridge from the age of two weeks.^29,30^ Meanwhile, in other cultures, there is a growing belief that babies who have grown for nine months in the womb are considered ready to eat and drink from birth.^31^ This practice is generally influenced by the perception that breastfeeding alone is not enough to make the baby feel full, so the mother fails to give exclusive breast milk for 6 months and encourages the feeding of formula to her baby.^32–34^ A similar perception was also found in a study in Oman, which reported that mothers who did not breastfeed exclusively were two to three times more likely to have concerns that their babies were at risk of malnutrition if they were only breastfed until six months of age.^25^

This study shows that areas with higher maternal education levels do not necessarily have higher exclusive breastfeeding coverage. In line with research in Brazil and Ethiopia, showing that maternal education has no significant relationship with exclusive breastfeeding.^22,35^ Theoretically, higher education is expected to increase maternal knowledge and awareness of the importance of exclusive breastfeeding. Some studies show that mothers who are highly educated will give more exclusive breastfeeding than mothers with low levels of education.^36–38^ These findings contrast with studies in Africa that reported higher exclusive breastfeeding coverage (48.0%) in communities with low levels of education than in higher-education communities (43.5%).^39^ This condition is allegedly associated with high work participation among educated mothers and exposure to formula milk marketing. However, the success of exclusive breastfeeding is determined not only by formal education but also by the understanding of breastfeeding acquired through counseling and education provided by health workers. Mothers with greater knowledge of breastfeeding are more likely to exclusively breastfeed.^20,28,40,41^ Compared with children in rural areas, those in urban areas are more likely to receive exclusive breastfeeding, likely due to greater access to information and health services.^36^

This study shows that areas with high proportions of facility-based deliveries do not necessarily have high exclusive breastfeeding coverage. Consistent with research conducted in Indonesia, there is no association between place of delivery and exclusive breastfeeding.^11^ These findings indicate that delivery in a health facility does not automatically guarantee the success of exclusive breastfeeding if it is not followed by ongoing postpartum support. Support from health workers is needed in breastfeeding counseling to promote understanding of the importance of exclusive breastfeeding.^42^ Research shows that breastfeeding mothers take twice as long to get support from midwives and hospital staff as mothers who don’t.^43^ Counseling and education carried out through health services and at the community level are the most effective interventions. Educating families and communities about breastfeeding and providing support to mothers may help create a better breastfeeding environment.^44^ The most important thing is that regular postpartum visits will allow mothers to get education about exclusive breastfeeding from health workers. Indonesian research indicates a significant association between healthcare providers and exclusive breastfeeding duration.^45^

In this context, barriers to successful breastfeeding may be influenced by limited support from health workers, infrequent lactation counseling, sociocultural factors, and family dynamics. Assistance by lactation consultants is essential, especially for mothers who experience the perceived insufficient milk supply (PIMS), through efforts to improve breastfeeding techniques, such as proper positioning and attachment, recommendations for breastfeeding on demand 8-12 times a day from both breasts, the application of breast compression during breastfeeding, and pumping breast milk after or between breastfeeding times.^44^ The findings showed that infants who received correct breastfeeding techniques through maternal intervention within 6 days of birth had fewer breastfeeding problems and a longer duration of breastfeeding than infants who received improper breastfeeding techniques.^32^ Thus, strengthening lactation counseling and breastfeeding assistance can support the success of exclusive breastfeeding.

## CONCLUSION

This study shows spatial variation in exclusive breastfeeding coverage in Muara Enim Regency, with only one village exhibiting relatively high coverage compared with other regions. These findings indicate interregional inequalities in the success of exclusive breastfeeding, influenced by social and health determinants, such as health worker support, access to lactation counseling, and the role of the family and social environment. The observed spatial patterns confirm that the success of exclusive breastfeeding is determined not only by individual maternal factors but also by the surrounding regional context and the support system. Therefore, a more targeted, region-based intervention is needed, particularly in villages with low coverage, by strengthening maternal and child health services, improving the quality of lactation counseling, and involving families and communities to increase exclusive breastfeeding coverage more evenly in Muara Enim Regency.

### Strengths

- Utilized a descriptive spatial approach to visualize geographic disparities in exclusive breastfeeding coverage.
- Based on aggregated individual-level questionnaire data from a relatively large sample (1,267 mothers).
- Applied spatial overlay and pie diagram mapping to display multiple variables across village and sub-district areas.

### Limitations

- Relied on secondary and self-reported data, which may affect data completeness and accuracy.
- Descriptive and area-level analysis did not allow causal inference.
- Data aggregation at village and sub-district levels may obscure within-area variation (ecological fallacy).

## Data Availability

All data produced in the present work are contained in the manuscript

## Funding

Not applicable.

## Ethics approval

This research has received approval from the Ethics Committee of the Faculty of Public Health, Sriwijaya University, with the ethics approval certificate number 748/UN9. FKM/TU. KKE/2025.

## Conflict of interest

There is no conflict of interest.

## Aknowledgment

We would like to express our gratitude to the Faculty of Public Health, Sriwijaya University for providing access to this data.

## Notes

### Competing Interest Statement

The authors have declared no competing interest.

### Funding Statement

This study did not receive any funding

